# Impact of healthcare worker shift scheduling on workforce preservation during the COVID-19 pandemic

**DOI:** 10.1101/2020.04.15.20061168

**Authors:** Dan M. Kluger, Yariv Aizenbud, Ariel Jaffe, Fabio Parisi, Lilach Aizenbud, Eyal Minsky-Fenick, Jonathan M. Kluger, Shelli Farhadian, Harriet M. Kluger, Yuval Kluger

## Abstract

**Background:** As we contend with the massive SARS-CoV-2 pandemic, preventing infections among healthcare workers (HCWs) and patients is critical for delivering care to patients admitted for other purposes, and many standard scheduling practices require reassessment. In most academic hospitals in the United States, inpatient rotations are designed to deliver optimal patient care by staggering rotations of attendings and house-staff, and much emphasis is placed on HCW burnout, yet during a pandemic preventing further infection is the single most important factor. Our purpose was to model various inpatient rotation schedules of physicians and nurses to determine patterns associated with optimal workforce preservation and lower nosocomial infections in settings in which personal protective equipment is imperfect or unavailable.

**Summary of Methods:** We simulated the spread of COVID-19 in hospital wards using Monte Carlo methods. Universal model parameters for COVID-19 included incubation period distribution and latent period distribution. Situation-dependent COVID-19 model parameters included pre-admission infection probability, team member infection probability, physician-to-patient, nurse-to-patient, patient-to-physician, patient-to-nurse, and HCW-to-HCW transmission probabilities, team member absence after symptom onset, daily SARS-CoV-2 exposure probability of team members (e.g. via exposure to other staff), length of admission after COVID-19 symptoms, and length of simulation time. Model parameters that varied by hospital setting and service type included average patient load per team, average patient hospitalization, and number of physicians and nurses on a team and on duty.

**Results:** The primary outcome measure was probability of team failure, defined as the likelihood that at some point there are insufficient attendings, house-staff or nurses to staff a fully functioning floor. In all of our simulations, physician and nurse rotation lengths of 1-3 days led to higher team failure rates. Nursing shifts of 12 versus 8 hours and avoiding staggering of physician rotations also decreased the chance of team failure.

**Conclusions:** Simple changes in staff scheduling, such as lengthening nursing shifts or avoiding rotations that are either staggered or last fewer than three days, can result in improved workforce preservation. These workforce scheduling changes are easy to implement.

---

As the COVID-19 pandemic continues, healthcare workers (HCWs) report for duty, caring for both COVID-19 patients and patients with non-COVID-19 conditions. Experiences in China and Italy suggest that HCWs are highly vulnerable to COVID-19 infection: in Italy, 20% of HCWs became infected with SARS-CoV-2 at the peak of disease spread.[1] Preventing COVID-19 infections among HCWs is critical for their safety and for stability of the healthcare delivery system. This includes stable functioning of non-COVID-19 wards, where HCWs may be exposed to SARS-CoV-2-infected patients who may not have undergone testing due to low clinical index of suspicion.

One way to reduce infection rates is to optimize staff scheduling to minimize interactions between different HCWs and limit the patient pool to which HCWs are exposed. Despite reports of nosocomial infections, infection of HCWs by patients, and transmission of SARS-CoV-2 from one HCW to another, little is known about the effects of HCW team structure on hospital transmission of SARS-CoV-2.[2, 3] Experience from other pandemics is not necessarily applicable, as infection and fatality rates are different. We therefore ran Monte Carlo simulations to explore various staffing possibilities with the goal of identifying staffing structures to minimize infections among HCWs on non-COVID-19 wards. For COVID-19 wards, in which the rate of patient-to-HCW transmission depends on personal protective equipment (PPE) and types of procedures and patient encounters, alternative input parameters for such simulations are needed; here we solely address staffing in non-COVID-19 wards.

For the five different scheduling designs represented in Figure 1, we simulated the spread of SARS-CoV-2 in hospital wards with various choices of model input parameters. Universal model parameters for COVID-19 included incubation period distribution (time from exposure to first symptom) and latent period distribution (time from exposure to becoming infectious.) Situation-dependent COVID-19 model parameters included pre-admission infection probability of an admitted patient, team member infection probability at start of simulation, physician-to-patient, nurse-to-patient, patient-to-physician, patient-to-nurse, and HCW-to-HCW transmission probabilities, team member days of absence after symptom onset, daily SARS-CoV-2 exposure probability of team members (e.g. via elevator use, exposure to other staff), length of patient stay after showing COVID-19 symptoms, and length of simulation time. Model parameters that varied by hospital setting and service type included average team patient census, average patient hospitalization length, and the number of physicians and nurses on a team and on duty at all times.

**Figure 1.**
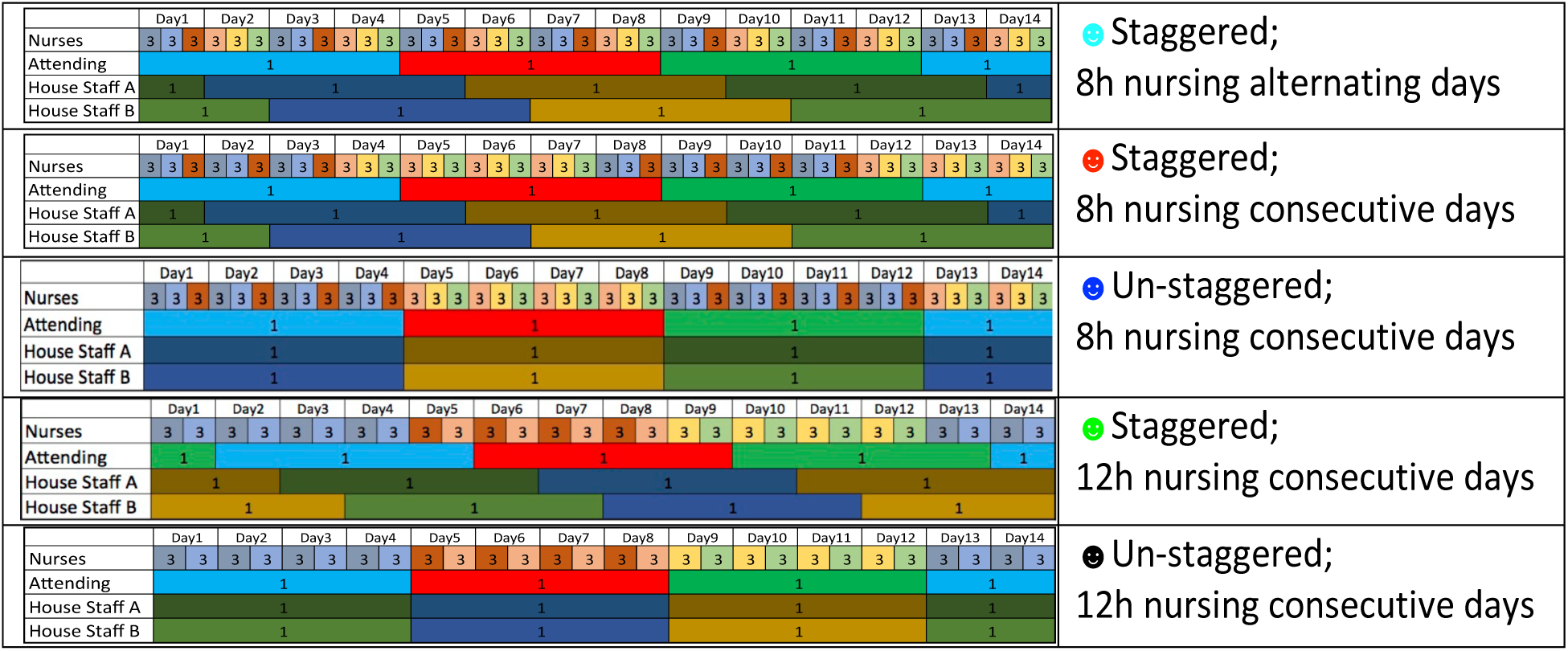
Scheduling designs. Schematic diagrams of five different scheduling designs for a team of 18 nurses, 3 attending physicians and 6 house-staff. These diagrams correspond to the scenario in which physicians rotate every four days, and when applicable, cohorts of nurses rotate every four days as well. Each physician is represented by a unique color. In each shift there are three nurses (triplet). The identity of the nurses in each triplet is fixed as long as all nurses in the triplet are healthy. Each triplet is represented by a unique color. The right column describes the different scheduling designs. The colors of the bullet points are matched with the colors representing the scheduling designs of Figure 2.

To illustrate how scheduling decisions affect infection rates, in Figure 2 we simulate two hospital teams, each including six house-staff or advanced practice providers (APPs) and three attending physicians, two house-staff/APPs and one attending on rotation at a time. The first team had 30 nurses (five per shift), and the second 18 nurses (three per shift.) The average number of patients is set to 15 per day (5 per nurse or 3 per nurse, in settings with different patient acuity.) Under normal circumstances, personnel rotations are staggered to ensure continuity of care and broad exposure for trainees to attendings and patients to enhance their educational experience. Rotation duration is also geared towards minimizing HCW fatigue. In a pandemic, these factors are considerably less important than HCW preservation. We compared scheduling options to minimize team failure, defined as the event that at some point there are insufficient attendings or house-staff/APPs to staff a fully functioning floor or insufficient healthy nurses to limit weekly hours to 48. Under all scenarios modeled each nurse works an average of ≤36 hours/week. Figure 1 illustrates five staff scheduling designs for a team of 18 nurses, 3 attending physicians and 6 house-staff with physician rotations duration of four days. Figure 2 depicts the outcomes of the five staff scheduling scenarios for a mean patient hospital stays of two and five days, typical for maternity and medicine floors, respectively, indicating team failure probability as a function of physician rotation length. We simulate situations in which cohorts of nurses co-rotate with physician rotations compared to nursing schedules that are independent of physician schedules.

**Figure 2.**
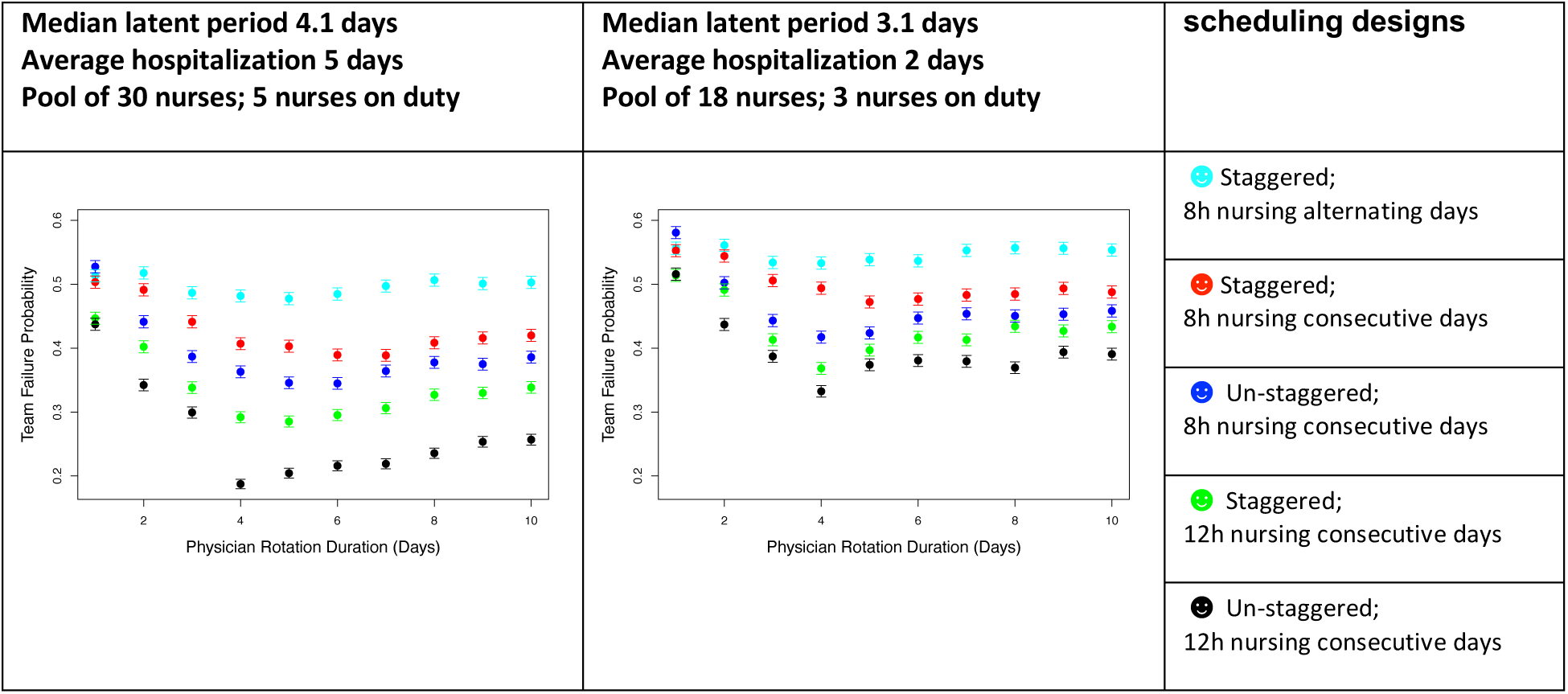
Probability of team failure vs. physician rotation duration. Team failure probability is based on Monte Carlo simulations plotted by duration of physician rotation, modeled for a team caring for patients with five-day average hospitalizations with fewer patients per nurse, such as internal medicine wards (left) or for patients with two-day average hospitalizations and more patients per nurse, such as maternity wards (right.) The plots compare the probability of team failure for five different scheduling designs. The designs simulated vary by whether they are staggered versus un-staggered, whether they have 8-hour nurse shifts versus 12-hour nurse shifts, and whether nurses work consecutive days versus work alternating days. Note that in our simulations with nurses working consecutive days, when the physician rotations are sufficiently short, the nurses work the same number of consecutive days as the physician do; however, if the physician rotations are too long, the nurses are scheduled to work as many consecutive days as possible without exceeding 48 hours of work in the span of one week, and without exceeding 36 hours/week on average. Of note, due to unknown variables in the model, these plots do not suggest that the actual probability of team failure lies in the 20-60% range, but rather the plots are intended to demonstrate the relative improvement of various staff scheduling changes. From the plots above, and from similar plots that we generated with varying choices of the unknown parameters, we observe that scheduling designs with un-staggered rotations, 12-hour nursing shifts over consecutive days are favorable, and further, the probability of team failure is lower when all HCWs work at least 3-4 consecutive days.

While the precise latent period of SARS-CoV-2 is unknown, the median incubation period is 5.1 days.[4] COVID-19 patients are likely most infectious 24 hours before and 24 hours after first symptoms.[5] Without frequent testing, shorter rotations increase the likelihood that infected HCWs will be off-rotation 24 hours before initiation of symptoms, while longer rotations expose fewer HCWs to the same infectious patient.

The rotation length that minimizes failure probability mainly depends on two factors: the median SARS-CoV-2 latent period, which is not precisely known, and the average hospitalization duration, and further understanding of the relationship between these factors is needed to make strong recommendations about optimal rotation length. However, in all simulations analyzed, physician and nurse rotation lengths of 1-3 days led to higher team failure rates; shorter rotations result in exposure of more HCWs to an infected patient. When the average patient stay is much longer than 5 days or when the median latent period is much shorter than 4 days, the benefit of un-staggering rotations decreases (data not shown). When patient stays are short, such as on maternity wards, the advantage of un-staggered rotations is consistent and universal across various parameters. Of note, because the actual probability of team failure is sensitive to other unknown parameters, plots such as those in Figure 2 should be used only to design optimal scheduling of shifts and not to forecast the actual probability of team failure.

In summary, pandemics necessitate widespread reassessment of workforce planning to ensure backup of sufficient uninfected HCWs. Using various input variables for our simulations for non-COVID-19 services, we make three primary observations: 1) Having all HCWs work at least three consecutive days reduces the chance of team failure, 2) longer nursing shifts (12 versus eight hours) decrease the rate of HCW infection, and 3) avoiding staggering of rotations of attendings, house-staff and nurses reduces the number of infected HCWs. When applying this model to the real-world challenge of staffing hospital units, clinical setting variables such as trainee presence, patient acuity, stay length, nursing/patient ratio, will need to be considered. Similar modeling can be employed for teams treating known COVID-19 patients.

In conclusion, alternative staffing methods, in which groups of physicians and nurses share rotations that are at least three days long with 12-hour nursing shifts, should be considered for workforce preservation in the COVID-19 pandemic.

## Data Availability

This study is not based on newly generated data

https://github.com/KlugerLab/Rotation-Scheduler

## Appendix/Methods

We designed a simulator that can run many repeated trials for any given set of input parameters. In each simulation, the medical team and patient system evolves for 180 days or until the team fails. Team failure is defined as the event that there are not enough healthy and available medics to staff a fully functioning team for a day. We deem nurses unavailable if they have worked at least 48 hours in the previous seven days.

On day zero in our simulations, each HCW and each of the initial 15 patients has a 0.001 probability to be infected with SARS-CoV-2. Further, each HCW and patient who was infected by the virus on day zero, was randomly assigned an incubation period t_inc_ from a lognormal distribution with median 5.1, and was randomly assigned a number from a Uniform(0, t_inc_) distribution to randomly determine how many days into the infection, each infected person is on day zero of the simulation.

The simulation progresses each day in multiple stages:

### First stage

We simulate the daily transmission of the virus between HCWs and patients. In particular, each of the physicians interacts with all of the patients. When a patient and a physician interact if the patient is infectious (a person is infectious beginning one or two days prior to the onset of their first symptom) and the physician has not yet been infected, the probability of patient to physician transmission was set to 0.1. Similarly, when a patient and a physician interact, and the physician is infectious, the probability of transmission from physician to uninfected patient was set to 0.1.

Patient-nurse interactions are subsequently simulated in the same manner, with two key differences. First, the patients are partitioned into k groups, where k is the number of nurses on a shift simultaneously, and each nurse only treats patients in one of the k groups. Because nurses spend more time than physicians do with each individual patient, the probability of a transmission between a nurse-patient pair that sees each other was set to a higher value than that for a physician-patient pair. Second, the probability of nurse to patient transmission and patient to nurse transmission is appropriately adjusted based on duration of the nursing shifts (for example, the probability of no transmission from an infectious patient to a nurse that works 8 hour shifts will differ by an exponent of 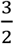 from that for a nurse that works 12 hour shifts.)

Transmissions between HCWs are then simulated. To do this we compute N_eff_, the effective number of infectious HCWs. N_eff_ is calculated based on how many infectious HCWs are present and the number of hours each of those infectious HCWs works. Then we simulate whether or not each individual healthy HCW is infected by other HCWs with probability that depends upon both N_eff_ and the number of hours that this healthy individual HCW works.

In the simulations we looked at, we assume that patients do not transmit the virus directly to other patients as all patients are in separate rooms for hospitals we simulate. Including an option to simulate transmission in hospital wards where there is more than one patient per room is simple and does not require restructuring.

Finally, for each day of work, additional sources of infections can be introduced, such as infections in elevators, short interactions with other staff such as pharmacists, clerical staff and janitors, and infections outside of the hospital such as family exposure and supermarket runs. These affect each HCW with a small probability (less than 0.0001 per day in our simulations).

Note that for all of the simulated transmissions mentioned above, transmission is only assumed to occur to HCWs and patients who have not yet had the virus, and immunity is assumed once a HCW recovers from the virus. Finally, once all of the above transmissions are simulated, each newly infected patient and each newly infected HCW is assigned an independently drawn incubation period from a lognormal distribution with a median of 5.1 days.

### Second stage

This stage of the simulation for the work day involves a random process for releasing and admitting new patients. After the daily transmissions are simulated, some patients will randomly leave and some new patients will be hospitalized. In particular, at the end of each day, each patient leaves with probability 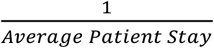, and on average 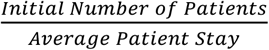 new patients are hospitalized at the end of each day. This method of randomly accepting and discharging patients, gives us the desired average patient stay (an input parameter for our model.) The method also gives us a consistent average number of patients present equal to the initial number of patients (also an input parameter for the model.) Finally, we assume that patients who have exhibited COVID-19 symptoms for at least 24 hours are identified, isolated, and no longer seen by physicians and nurses on the team in person.

### Third stage

This stage of the simulation determines at each day which HCWs will work the following day. At the end of each day in the simulation (midnight), any HCW who began showing COVID-19 symptoms 21 days prior is either put in the category of “very ill” with probability 0.07 after which this HCW is unable to return to work, or is put in the category of “recovered” after which the HCW is able to return to work and is immune to the virus. In addition, at midnight, each HCW who started showing COVID-19 symptoms that day is replaced by the healthy HCW in the same category who has been waiting at home for the longest period of time. Further each nurse who has worked 48 or more hours in the past seven days is replaced by the nurse who has been off duty for the longest period of time among healthy nurses. If at least one of these replacements cannot be made, the team fails and the simulation ends.

In addition to replacing HCWs who start showing COVID-19 symptoms, physician and nursing rotations are also implemented in stage three. We have input parameters T_Physician_ and T_nurse_ to denote the number of consecutive days each physician and nurse is scheduled to work respectively. In the staggered setting, each HCW is replaced, whenever possible, after a physician works T_Physician_ days and after a nurse works T_nurse_ days. In the un-staggered setting, after every T_nurse_ days, all nurses who were on duty and can be replaced are replaced, and after every T_Physician_ days, all physicians who are on duty and can be replaced are replaced.

After substituting symptomatic HCWs and fulfilling HCW rotations, the simulation progresses to the next day. The days in simulation progress until either the team fails or the 180 days completed without team failure.

So far, we described a single simulation. To estimate team failure probability for a given set of parameters and schedule design, we execute 10,000 simulations with same parameter values. Team failure probability is estimated by computing the proportion of simulations that result in team failure.

Rotation-Scheduler was programmed in R. Rotation-Scheduler is available at https://github.com/KlugerLab/RotationScheduler. The code for all experiments is available on request and will be publicly available together with a detailed tutorial at https://github.com/KlugerLab/Rotation-Scheduler-paper on publication.

## Notes

### Competing Interest Statement

The authors have declared no competing interest.

### Funding Statement

None

